# Air pollution drives concentrated clinical severity of respiratory illness in early childhood

**DOI:** 10.64898/2026.09.03.26362213

**Authors:** Haedong Kim, Junghwan Suh, Jaeyong Shin, Hee-Seung Yang

**Affiliations:** Department of Preventive Medicine, Yonsei University College of Medicine, Seoul, Republic of Korea; Department of Paediatrics, Severance Children’s Hospital, Yonsei University College of Medicine, Seoul, Republic of Korea; School of Economics, Yonsei University, Seoul, Republic of Korea

## Abstract

Exposure to fine particulate matter (PM_2.5_) increases paediatric respiratory morbidity, but its impact on the intensity and economic burden of care in early childhood remains unclear. Here we link nationwide health records for 3,210,381 South Korean children (99,261,945 child-month observations, 2015–2019) and exploit quasi-random variation in PM_2.5_ induced by thermal inversions to estimate its causal effects on respiratory care and expenditure. A 1-standard-deviation increase in monthly PM_2.5_ (7.23 µg/m³) raises respiratory visits broadly, peaking at age 1 (0.29 additional visits per child-month) and remaining significant through age 6. In contrast, expenditures rise disproportionately among children under 3, increasing by 46.7%, 43.0%, and 35.4% at ages 0, 1, and 2 and becoming indistinguishable from zero beyond age 4. This shift towards higher-intensity emergency care translates into a 32.2% increase in annual respiratory spending among children under 5. Air pollution thus imposes a dual burden, amplifying both the frequency and severity of illness among the youngest infants.

---

Ambient air pollution is a major global health challenge, and among air pollutants, fine particulate matter (PM_2.5_) imposes the largest health burden, contributing to millions of premature deaths and substantial losses in human capital annually^1^. According to the Global Burden of Disease Study, this exposure is a leading driver of lower respiratory infections and chronic lung disease, with a disproportionate impact on children^2^. Quasi-experimental studies show that sharp reductions in pollution, triggered by economic downturns, regulatory interventions, or industrial disruptions, improve infant survival and neonatal health across settings^3–14^. These studies establish that clean air is critical for early-life health.

Beyond the immediate clinical burden, the global economic implications of early-life pollution exposure are significant. Human capital formation is a cumulative process, and health shocks during critical developmental periods can have lasting effects on cognitive development, educational attainment, and future labour-market outcomes^15,16^. While the long-term scarring effects of pollution are increasingly recognised, the immediate channels through which these costs materialise, specifically the translation of ambient shocks into acute healthcare consumption, remain under-studied in the paediatric population. Understanding this translation is crucial for healthcare budgeting and for identifying the most vulnerable developmental windows.

Clinical and epidemiological evidence shows that even a modest deterioration in air quality can trigger wheezing and asthma exacerbations and increase paediatric respiratory morbidity^17,18^. Global assessments attribute a significant fraction of paediatric asthma incidence to ambient pollution^19,20^, which drives acute asthma-related symptoms and utilisation^21,22^. Physiologically, this is unsurprising; in early life, airway calibre is small, alveoli are still forming, and immune surveillance is incomplete^23,24^. PM_2.5_ can also activate innate immune pathways and alter airway macrophage function, intensifying inflammation during acute respiratory insults^25–28^.

Despite this biological vulnerability, most economic studies of pollution-related medical costs have focused on adult and older populations^29–31^. For children, existing studies often aggregate those under age 5 due to data constraints, risking the masking of important developmental heterogeneity. If a 6-month-old infant and a 4-year-old preschooler are treated as equivalent, specific windows of vulnerability, and the associated ‘hidden costs’, may be overlooked. Consequently, this study provides population-scale, age-resolved causal evidence on how PM_2.5_ affects early-childhood respiratory healthcare use and spending in a universal-coverage setting. Leveraging thermal inversions as an instrumental variable, we separate the morbidity margin from the expenditure margin to identify where transient particulate shocks impose the largest healthcare burden along the age distribution. Here we show that transient PM_2.5_ shocks broadly increase respiratory visits across early childhood yet concentrate clinical severity and cost among children under three, revealing a divergence between how often children are seen and how severely they are treated. As these developmental vulnerabilities are biologically universal and the identification strategy is portable to other settings with administrative health data, the findings carry implications well beyond Korea for regions facing high particulate exposure.

## Results

### A natural laboratory

We studied South Korea because it is a high-income universal-coverage setting that combines recurrent PM_2.5_ episodes with near-census administrative health data. As of 2019, South Korea recorded the highest annual PM_2.5_ exposure among Organisation for Economic Co-operation and Development countries, exceeding other member states by a substantial margin^1,32^. Particulate pollution reflects a mix of local emissions from industry, traffic, power generation, and residential heating, together with regional transport in East Asia^33–35^. Exposure varies markedly by season; stagnation and heating demand increase concentrations in winter and early spring, whereas monsoon rainfall and enhanced vertical mixing reduce them in summer (Fig. 1). While informative, these cycles do not provide credible identification, because winter coincides with the peak transmission of influenza and respiratory syncytial viruses.

**Fig. 1.**
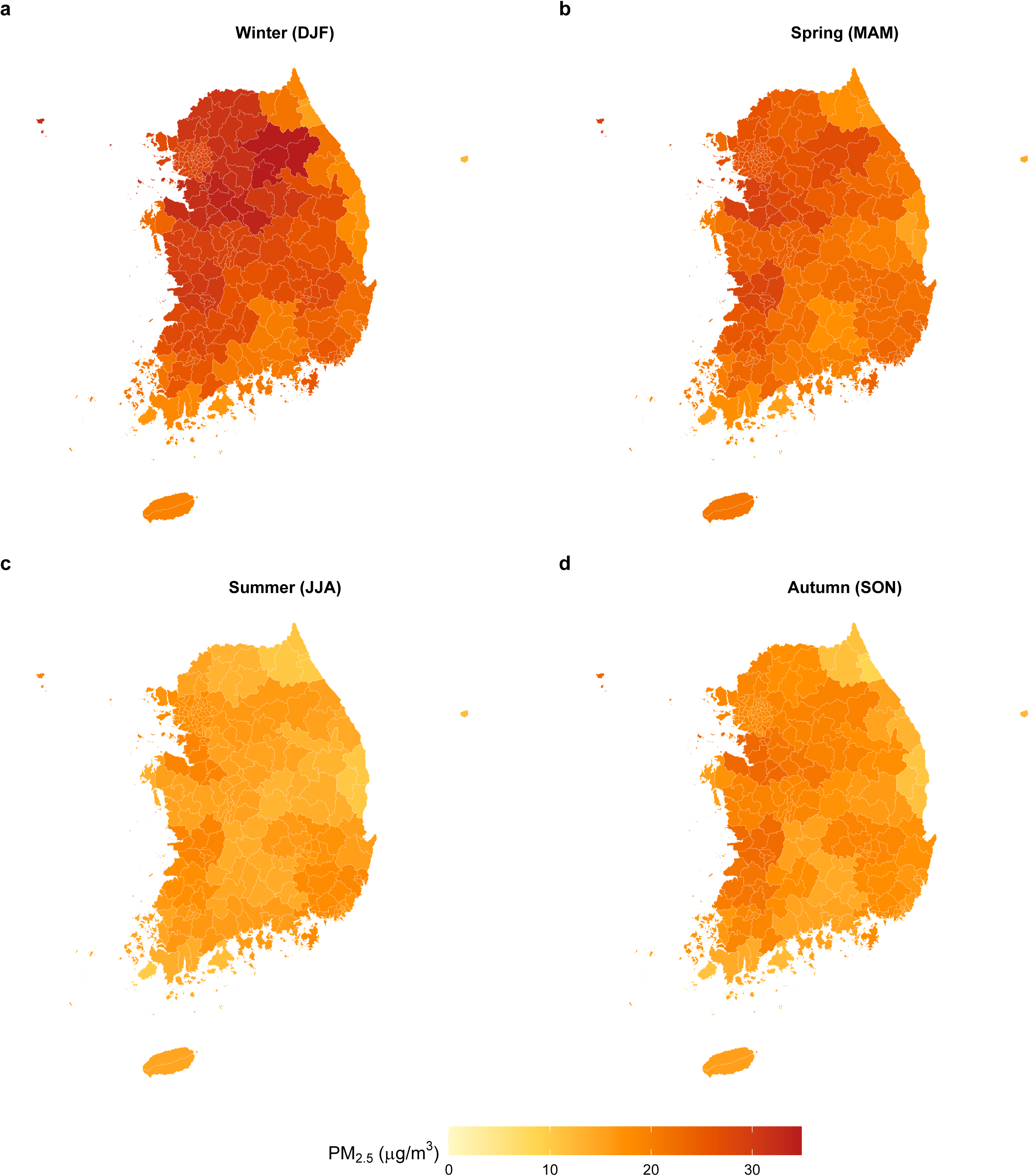
Seasonal patterns of ambient PM_2.5_ across South Korea (2015–2023). **a** Winter (December–February). **b** Spring (March–May). **c** Summer (June–August). **d** Autumn (September–November). Winter and early spring exhibit persistently high concentrations, especially in inland basins and downwind of major urban–industrial clusters, whereas summer and autumn show comparatively lower levels. PM_2.5_ concentrations are from the National Institute of Environmental Research. NIER National Institute of Environmental Research, PM_2.5_ particulate matter with aerodynamic diameter ≤2.5 µm.

South Korea’s health data infrastructure provides near-complete observability of paediatric care. Universal health insurance generates electronic claims for outpatient visits, emergency attendance, and hospital admissions with detailed payment components. Linking these claims to child-screening programmes allows age to be measured in months and supports rich fixed-effects designs. Our final cohort comprised 3,210,381 children contributing 99,261,945 child-month observations between 2015 and 2019, with a mean monthly PM_2.5_ of 25.36 µg/m³ (SD 7.23), mean respiratory visits of 1.71 per child-month, and mean monthly respiratory expenditure of US$26.99 per child (Table 1). Thermal-inversion strength co-moves with PM_2.5_ (r = 0.45), supporting instrument relevance (Fig. 2), and respiratory spending co-moves closely with PM_2.5_; non-respiratory categories remain relatively stable (Fig. 3). This combination of high exposure variability and comprehensive healthcare data offers a transparent setting for tracing how short-term pollution shocks translate into paediatric healthcare use and clinical severity.

**Fig. 2.**
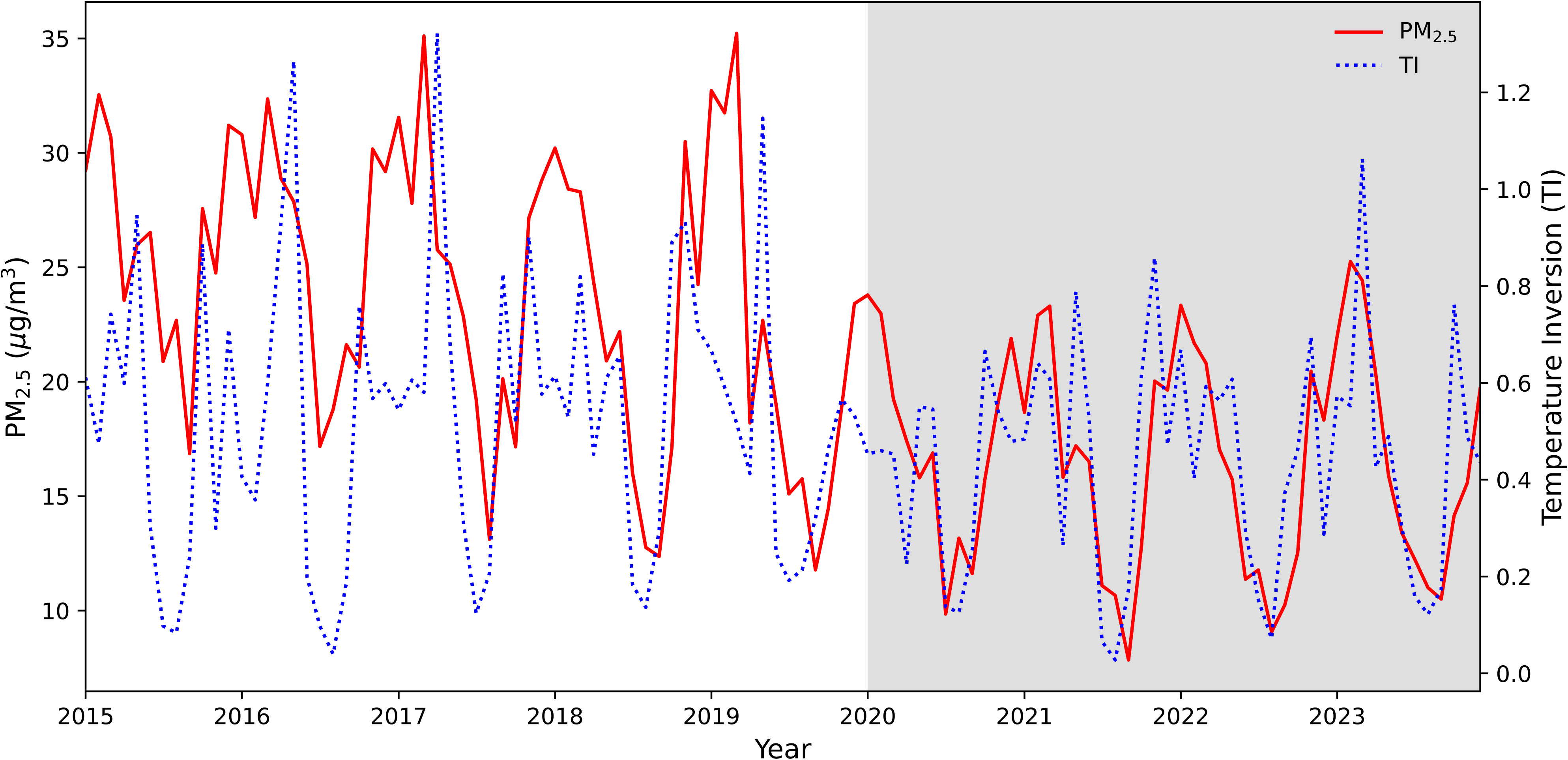
Co-movement of national monthly PM_2.5_ and thermal inversions. Solid and dashed lines plot national monthly PM_2.5_ and the thermal inversion index from 2015 to 2023, respectively. Peaks in inversion strength tend to coincide with peaks in PM_2.5_ (r = 0.45), consistent with inversions suppressing vertical mixing and trapping particulates near the surface. This co-movement indicates that inversions generate coherent exposure shocks at the national scale and supports their use as quasi-experimental shifters of local PM_2.5_ in the instrumental-variable analysis. Grey shading marks the post-2019 period (2020–2023), during which PM_2.5_ levels attenuated following the COVID-19 pandemic. Consequently, our primary analysis is restricted to the pre-pandemic period (2015–2019) to ensure estimate stability. PM_2.5_ concentrations are from NIER; thermal inversion strength is from NASA MERRA-2 reanalysis (M2I6NPANA, version 5.12.4). NIER National Institute of Environmental Research, TI thermal inversion.

**Fig. 3.**
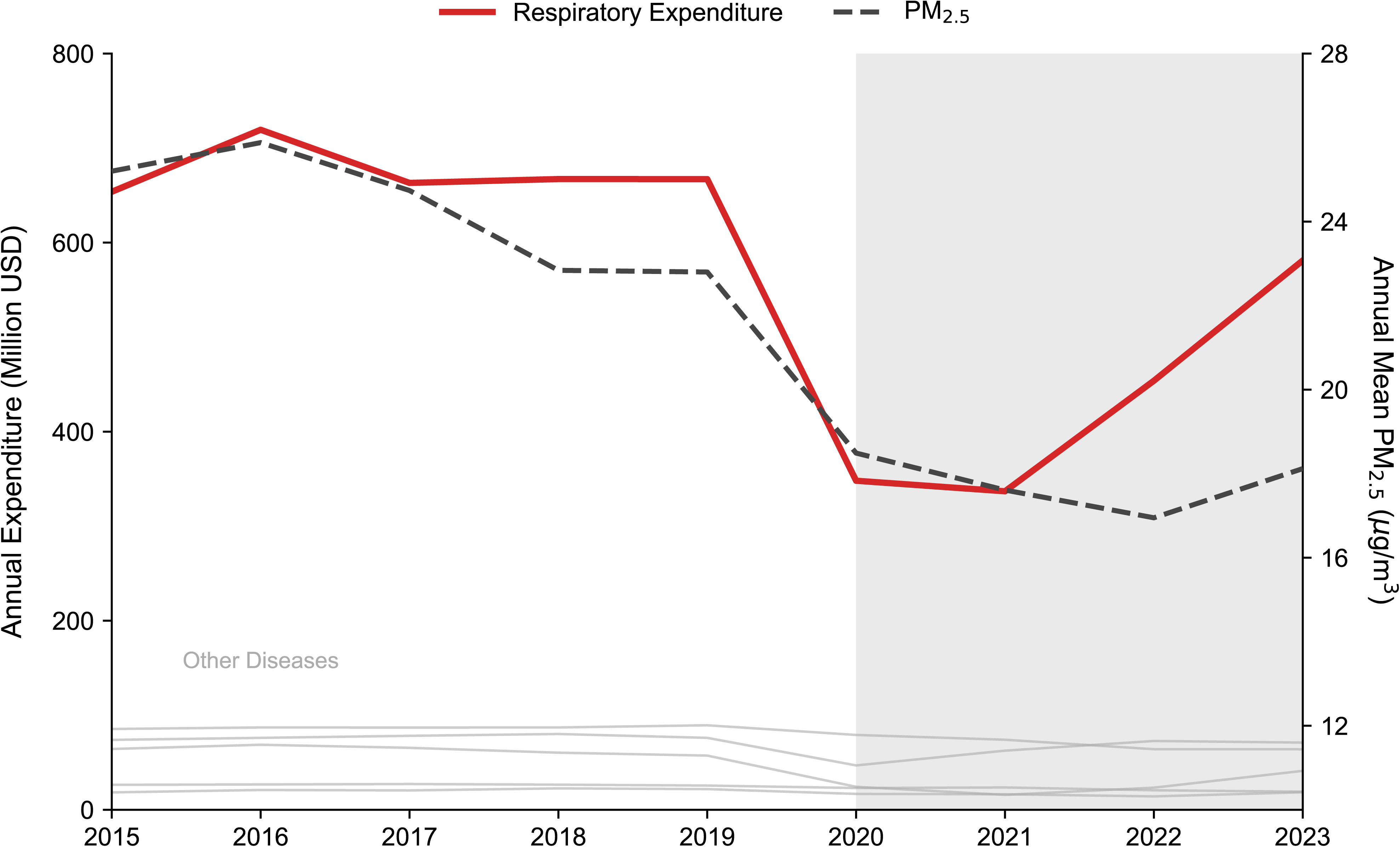
Healthcare expenditure for children under age 5 and PM_2.5_. Time series of annual healthcare expenditure for children under age 5 by broad diagnostic category, alongside national yearly PM_2.5_ from 2015 to 2023. Respiratory conditions dominate paediatric spending and co-move closely with PM_2.5_, whereas non-respiratory categories such as surgical and ophthalmologic care evolve more smoothly over time. In 2019, children aged 0–4 incurred US$667 million in respiratory healthcare expenditure; given that a 1-SD increase in monthly PM_2.5_ (7.23 µg/m³; Table 2) implies a 32.2% rise in spending, this corresponds to approximately US$215 million in excess costs. Grey shading marks the post-2019 period (2020–2023), during which PM_2.5_ levels and healthcare utilisation patterns were disrupted by the COVID-19 pandemic; consequently, our primary analysis is restricted to the pre-pandemic period (2015–2019) to ensure estimate stability. Data on healthcare expenditure are sourced from the Health Insurance Review and Assessment Service Open Data Portal. HIRA Health Insurance Review and Assessment Service, SD standard deviation.

**Table 1.** Summary statistic.

| Variable | Full sample (N=3,210,381) |
| --- | --- |
| **Health outcomes (monthly, per child)** |  |
| Respiratory visits | 1.71 (2.09) |
| Respiratory expenditure (US\$) | 26.99 (113.65) |
| Health insurance dues (US\$) | 97.07 (69.25) |
| **Environmental variables (monthly)** |  |
| PM<sub>2.5</sub> (µg/m<sup>3</sup>) | 25.36 (7.23) |
| Thermal inversion strength (°C) | 0.47 (0.82) |
| Precipitation (cm) | 7.31 (5.87) |
| **Child and household characteristics** |  |
| Age (months) | 48.06 (27.29) |
| Male sex | 1,650,786 (51.4%) |
| Disability status | 20,172 (<1%) |
| **Parental occupation** |  |
| Agriculture | 11,950 (0.4%) |
| Manufacturing | 1,279,431 (39.9%) |
| Retail | 653,448 (20.4%) |
| Services | 1,198,434 (37.3%) |
| Others | 67,118 (2.1%) |
| Child-month observations | 99,261,945 |

Notes: The sample covers 2015–2019 and comprises n = 3,210,381 children contributing 99,261,945 child-month observations. Data are n (%) or mean (SD). All variables were aggregated at the monthly level; PM<sub>2.5</sub> concentrations are available for 96,918,404 child- month observations. Health utilisation and payment data were obtained from the National Health Insurance Service (NHIS); PM<sub>2.5</sub> concentrations from the National Institute of Environmental Research (NIER); precipitation from the Korea Meteorological Administration (KMA); and thermal inversion strength was calculated using NASA’s MERRA-2 reanalysis products (M2I6NPANA, version 5.12.4). SD standard deviation.

**Table 2.**
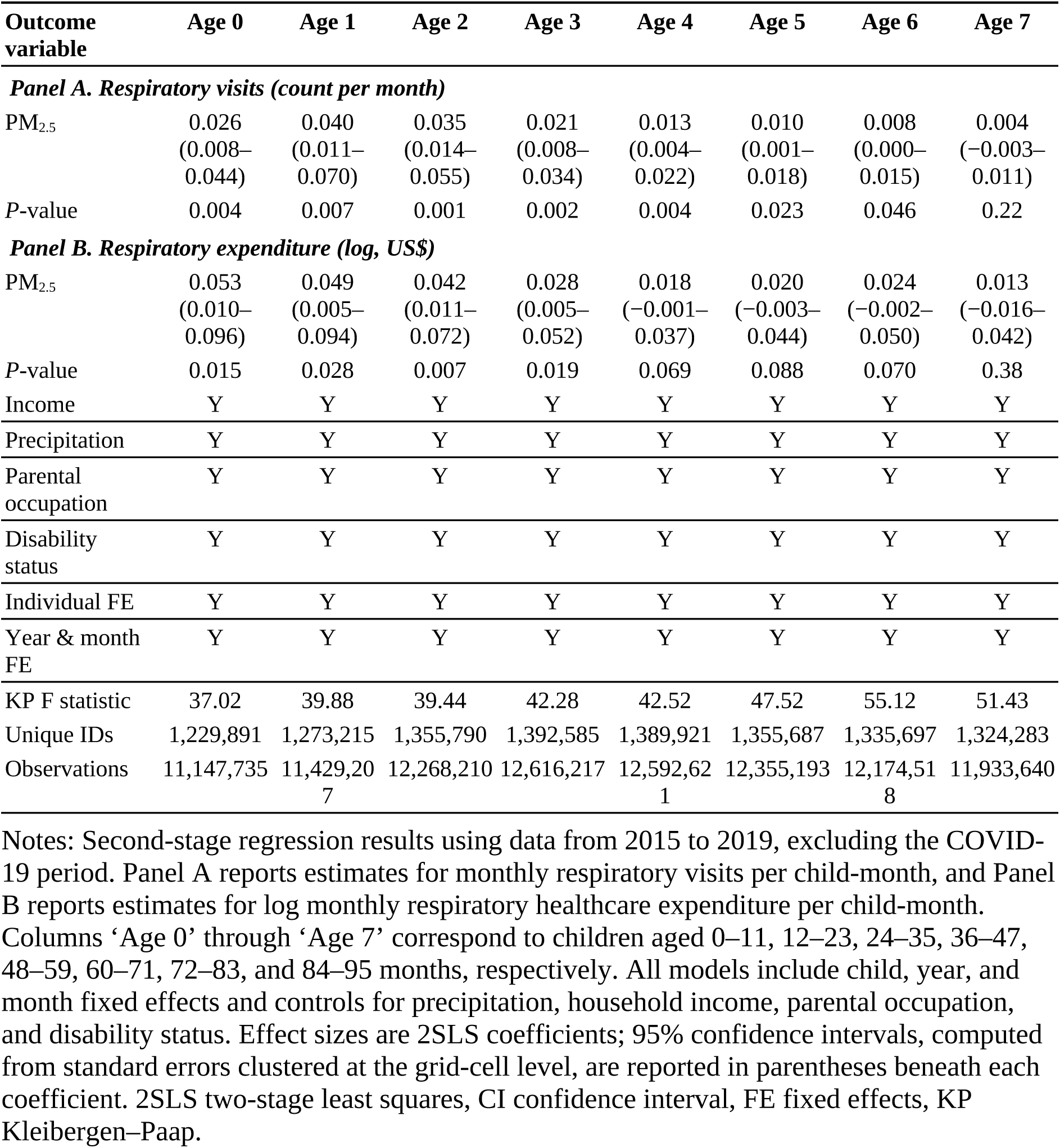
Age-specific causal effects of PM_2.5_ on respiratory outcomes (2015–2019)

| Outcome variable | Age 0 | Age 1 | Age 2 | Age 3 | Age 4 | Age 5 | Age 6 | Age 7 |
| --- | --- | --- | --- | --- | --- | --- | --- | --- |
| **Panel A. Respiratory visits (count per month)** |  |  |  |  |  |  |  |  |
| PM<sub>2.5</sub> | 0.026<br>(0.008–<br>0.044) | 0.040<br>(0.011–<br>0.070) | 0.035<br>(0.014–<br>0.055) | 0.021<br>(0.008–<br>0.034) | 0.013<br>(0.004–<br>0.022) | 0.010<br>(0.001–<br>0.018) | 0.008<br>(0.000–<br>0.015) | 0.004<br>(–0.003–<br>0.011) |
| P-value | 0.004 | 0.007 | 0.001 | 0.002 | 0.004 | 0.023 | 0.046 | 0.22 |
| **Panel B. Respiratory expenditure (log, US\$)** | | | | | | | | |
| PM<sub>2.5</sub> | 0.053<br>(0.010–<br>0.096) | 0.049<br>(0.005–<br>0.094) | 0.042<br>(0.011–<br>0.072) | 0.028<br>(0.005–<br>0.052) | 0.018<br>(–0.001–<br>0.037) | 0.020<br>(–0.003–<br>0.044) | 0.024<br>(–0.002–<br>0.050) | 0.013<br>(–0.016–<br>0.042) |
| P-value | 0.015 | 0.028 | 0.007 | 0.019 | 0.069 | 0.088 | 0.070 | 0.38 |
| Income | Y | Y | Y | Y | Y | Y | Y | Y |
| Precipitation | Y | Y | Y | Y | Y | Y | Y | Y |
| Parental occupation | Y | Y | Y | Y | Y | Y | Y | Y |
| Disability status | Y | Y | Y | Y | Y | Y | Y | Y |
| Individual FE | Y | Y | Y | Y | Y | Y | Y | Y |
| Year & month FE | Y | Y | Y | Y | Y | Y | Y | Y |
| KP F statistic | 37.02 | 39.88 | 39.44 | 42.28 | 42.52 | 47.52 | 55.12 | 51.43 |
| Unique IDs | 1,229,891 | 1,273,215 | 1,355,790 | 1,392,585 | 1,389,921 | 1,355,687 | 1,335,697 | 1,324,283 |
| Observations | 11,147,735 | 11,429,207 | 12,268,210 | 12,616,217 | 12,592,621 | 12,355,193 | 12,174,518 | 11,933,640 |

Notes: Second-stage regression results using data from 2015 to 2019, excluding the COVID-19 period. Panel A reports estimates for monthly respiratory visits per child-month, and Panel B reports estimates for log monthly respiratory healthcare expenditure per child-month. Columns ‘Age 0’ through ‘Age 7’ correspond to children aged 0–11, 12–23, 24–35, 36–47, 48–59, 60–71, 72–83, and 84–95 months, respectively. All models include child, year, and month fixed effects and controls for precipitation, household income, parental occupation, and disability status. Effect sizes are 2SLS coefficients; 95% confidence intervals, computed from standard errors clustered at the grid-cell level, are reported in parentheses beneath each coefficient. 2SLS two-stage least squares, CI confidence interval, FE fixed effects, KP Kleibergen–Paap.

### Identifying causal effects

Identifying the health effects of pollution is complicated because air quality is correlated with several other factors^4,6^. In Korea, winter and early spring bring higher PM_2.5_ alongside viral epidemics, school terms, and changes in indoor crowding; thus, simple comparisons between polluted and clean months would confound the toxicity of particulates with seasonal cycles in infection and behaviour. To address this, we exploited thermal inversions as quasi-experimental shocks to local PM_2.5_. In a standard atmosphere, warm air near the surface rises and disperses pollutants. During inversion episodes, a layer of warmer air overlies cooler surface air, suppressing vertical mixing and allowing pollutants to accumulate near the ground.

A key advantage of this instrument is the breadth of exposure it captures. Unlike the westerly-wind instruments used in previous Korean studies^32,36,37^, which capture only imported transboundary pollution, thermal inversions reflect the full local particulate load irrespective of prevailing winds, shifting the entire mixture of pollutants that children actually breathe (see Methods).

This approach isolates exogenous variation in PM_2.5_ from seasonal confounders, allowing us to distinguish the biological effects of pollution from spurious correlations driven by viral cycles or behavioural patterns, and is supported by null estimates in non-respiratory placebo tests. The robust physical linkage between inversions and particulate concentrations ensures that our estimates capture variation driven by meteorology rather than unobserved determinants of healthcare use^7,8,30^. First-stage instrument strength was strong throughout, with Kleibergen–Paap F statistics exceeding 37 in all age groups (Table 2). Details of the data sources, cohort construction, outcome definitions, and estimation are provided in the Methods section.

### Broad morbidity across childhood

We first examined the morbidity margin (respiratory encounters per child per month) to capture the extensive response to pollution (Fig. 4a). A 1-standard-deviation increase in monthly PM_2.5_ (7.23 µg/m³) significantly increased respiratory encounters across ages 0–6 years. The response peaked at age 1, inducing 0.29 additional visits per month, a noticeable increase relative to the sample mean of 1.71 visits (Table 1). Although this effect attenuated with age, consistent with the maturation of respiratory defence mechanisms, the effect remained robustly positive through age 6, implying that developing lungs remain sensitive to particulate irritation. Pollution thus imposes a widespread burden on the healthcare system by triggering symptoms that require medical attention.

**Fig. 4.**
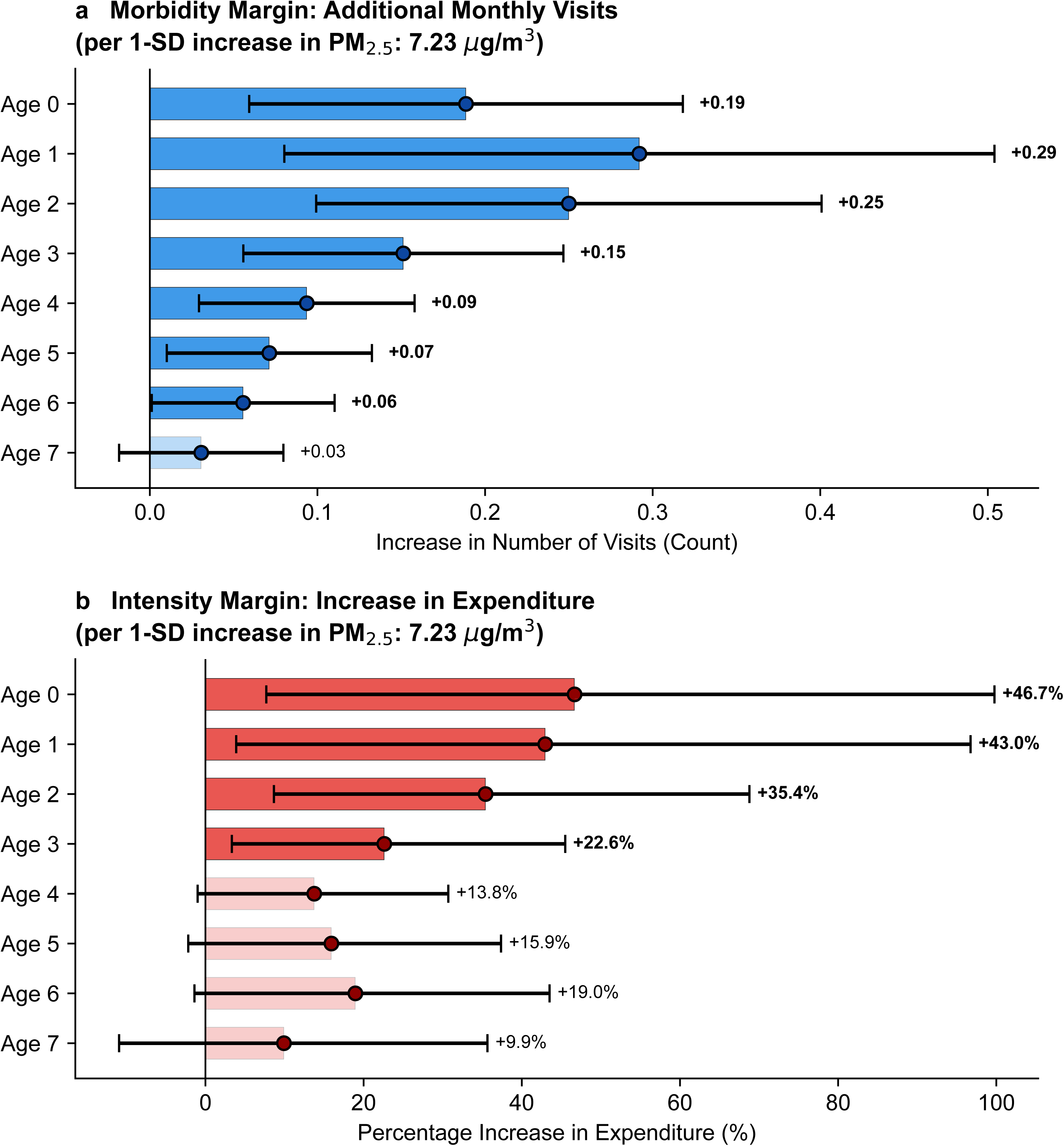
Age-specific effects of PM_2.5_ on respiratory visits and expenditure. **a** Instrumental-variable estimates of the effect of a 1-SD increase in monthly PM_2.5_ (7.23 µg/m³) on respiratory visits per child-month at ages 0–7 (n = 3,210,381). **b** Corresponding instrumental-variable estimates for monthly respiratory expenditure per child, including months with zero spending. Symbols: age-specific point estimates; horizontal lines: 95% CIs. Estimates and 95% CIs were obtained from 2SLS regressions instrumented by thermal inversions, adjusted for child, year, and month fixed effects and for precipitation, household income, parental occupation, and disability status. Visit responses remain positive well into early school age, whereas expenditure responses are sharply front-loaded: respiratory spending increases by 46.7% at age 0, 43.0% at age 1, 35.4% at age 2, and 22.6% at age 3, with smaller and imprecise effects thereafter. Thus, transient PM_2.5_ shocks generate broad paediatric respiratory morbidity, while financially severe episodes are concentrated in the first 3 years of life. First-stage Kleibergen–Paap F statistics consistently exceed 37 (Table 2). 2SLS two-stage least squares, CI confidence interval, SD standard deviation.

However, the trajectory at the beginning of life deviates from this linear decline. The response for infants at age 0 corresponds to 0.19 additional visits per month, lower than the peak of 0.29 observed among 1-year-olds. Within the first year of life, however, two distinct phases emerged (Fig. 5): the healthcare response was statistically indistinguishable from zero for newborns and early infants (0–5 months), consistent with a strong behavioural shielding mechanism whereby young infants are kept indoors. This pattern is more consistent with an ambient-pollution pathway than with seasonal viral confounders. Unlike air pollution, viral epidemics penetrate households via family transmission and may even affect homebound newborns; thus, the absence of effects in this group points to outdoor exposure as the driver. In contrast, for older infants (6–11 months), the response surged nearly five-fold relative to the first half of infancy. This sharp increase coincides with greater mobility and outdoor exposure. The contrast between 0.19 additional visits at age 0 and 0.29 at age 1 therefore should not be read as newborns being biologically resilient. Rather, the age-0 estimate is a weighted average of two developmentally distinct regimes: a null response over the first 6 months, when infants are largely shielded indoors and ambient particulates rarely reach them, and a steep response thereafter, once infants begin to move, are carried outdoors, and encounter ambient air directly. Biological vulnerability is present from birth; what changes at around 6 months is exposure. The 6-month transition does not mark the onset of susceptibility but the point at which susceptibility is finally met by exposure and begins to translate into measurable healthcare use and cost.

**Fig. 5.**
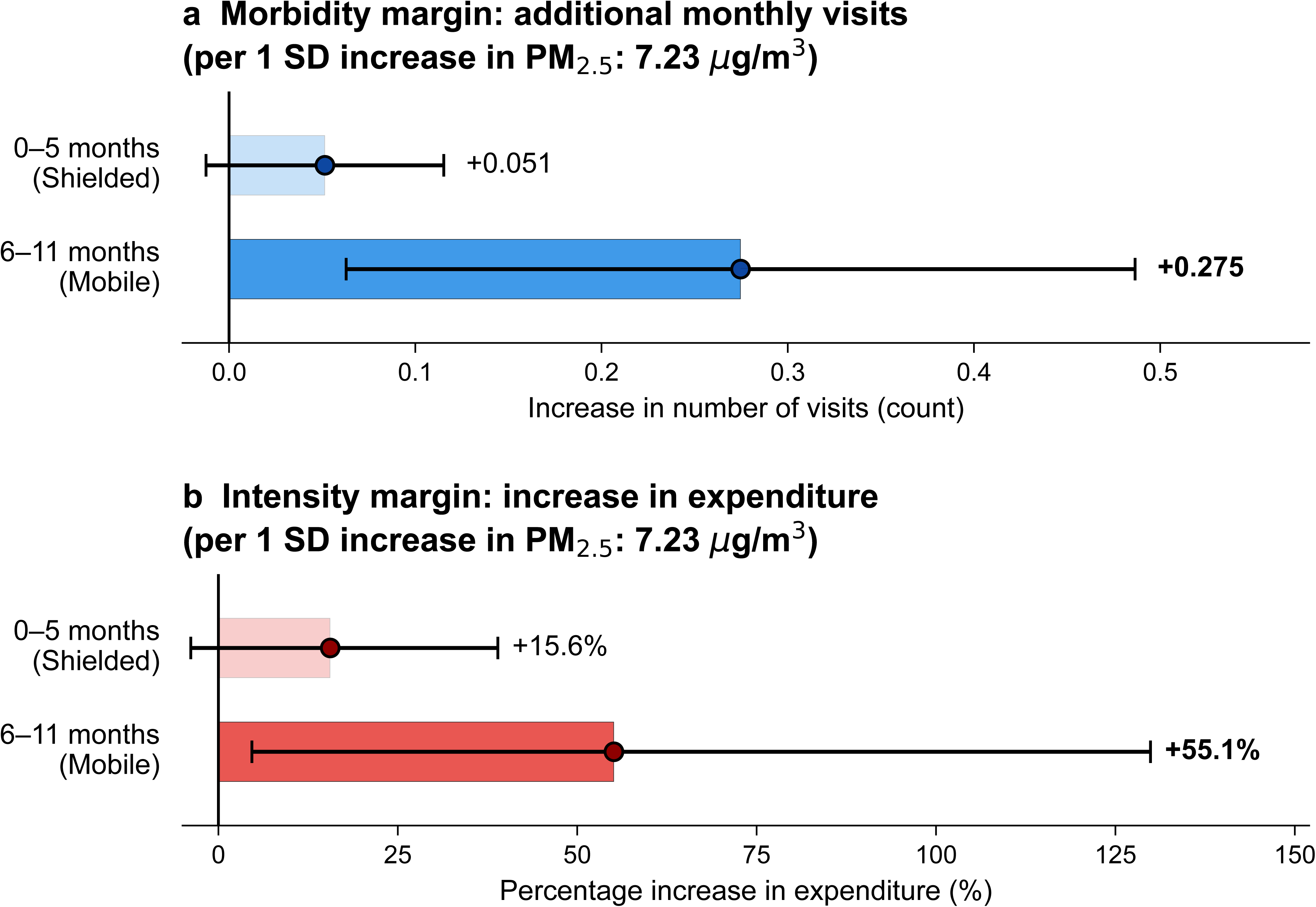
Heterogeneity in respiratory response during the first year of life. **a** Estimates of the effect of a 1-SD increase in monthly PM_2.5_ (7.23 µg/m³) on respiratory visits for infants aged 0–5 months (newborns and early infants) versus 6–11 months (older infants). The response is statistically insignificant in the first 6 months but increases sharply and becomes significant in the second half of infancy. **b** Corresponding estimates for monthly respiratory expenditure per child. Like visits, the financial burden is disproportionately concentrated at 6–11 months. This pattern is consistent with a behavioural shielding mechanism, where limited outdoor exposure in early infancy dampens the health shock, while increased mobility and outdoor activity after 6 months amplify it. Symbols: point estimates; error bars: 95% CIs. CI confidence interval, SD standard deviation.

We also examined biological susceptibility by stratifying analyses by sex. Results were broadly consistent across sexes, with both boys and girls showing significant morbidity responses of comparable magnitude across the high-risk window of 0–2 years (Supplementary Tables 1 and 2). For example, the effect on 1-year-olds is nearly identical for boys (coefficient = 0.041) and girls (coefficient = 0.040), suggesting that the pollution burden is a universal challenge for the developing paediatric respiratory system, regardless of sex.

### Concentrated costs in infancy

In sharp contrast to the broad morbidity response, the expenditure margin (monthly respiratory expenditure per child) is highly concentrated in early life (Table 2 and Fig. 4b). A 1-standard-deviation increase in PM_2.5_ increased respiratory expenditure by 46.7% for infants at age 0, with similarly large effects at ages 1 (43.0%) and 2 (35.4%). These estimates imply that for the youngest children, pollution does not merely cause more visits; rather, pollution fundamentally shifts the nature of care towards more expensive, resource-intensive interventions.

After age 3, the expenditure response declines markedly, falling to 22.6% at age 3 and becoming statistically indistinguishable from zero beyond age 4. This pattern reveals a clear developmental wedge between how often children are seen and how severely they are treated. Among older children, pollution continues to generate additional respiratory contacts, yet those contacts resolve as brief, low-cost outpatient encounters: the quantity of care rises while its intensity does not. In infancy, the same ambient shock produces a qualitatively different response, in which each additional encounter is more likely to require intensive, high-cost management; thus, total expenditure rises far more than visit counts alone would predict. The two margins therefore diverge with age: the morbidity margin extends broadly across childhood, whereas the expenditure margin closes sharply after age 3. A metric based on utilisation counts would register these two regimes as similar, obscuring precisely the ages at which pollution is most costly.

The concentration of excess expenditure in the first 3 years indicates an escalation in clinical severity. To identify the drivers of these costs, we distinguished routine outpatient care from emergency department (ED) encounters (Fig. 6). Decomposing expenditures by care setting revealed that outpatient spending increases with PM_2.5_, peaking at age 1 and declining thereafter, with age 0 remaining lower owing to behavioural shielding. By contrast, ED expenditures exhibit no such shielding and increase sharply for children aged 0–1, specifically at high PM_2.5_ in the highest deciles (8–10). This pattern indicates that pollution disproportionately induces episodes of higher acuity in early life.

**Fig. 6.**
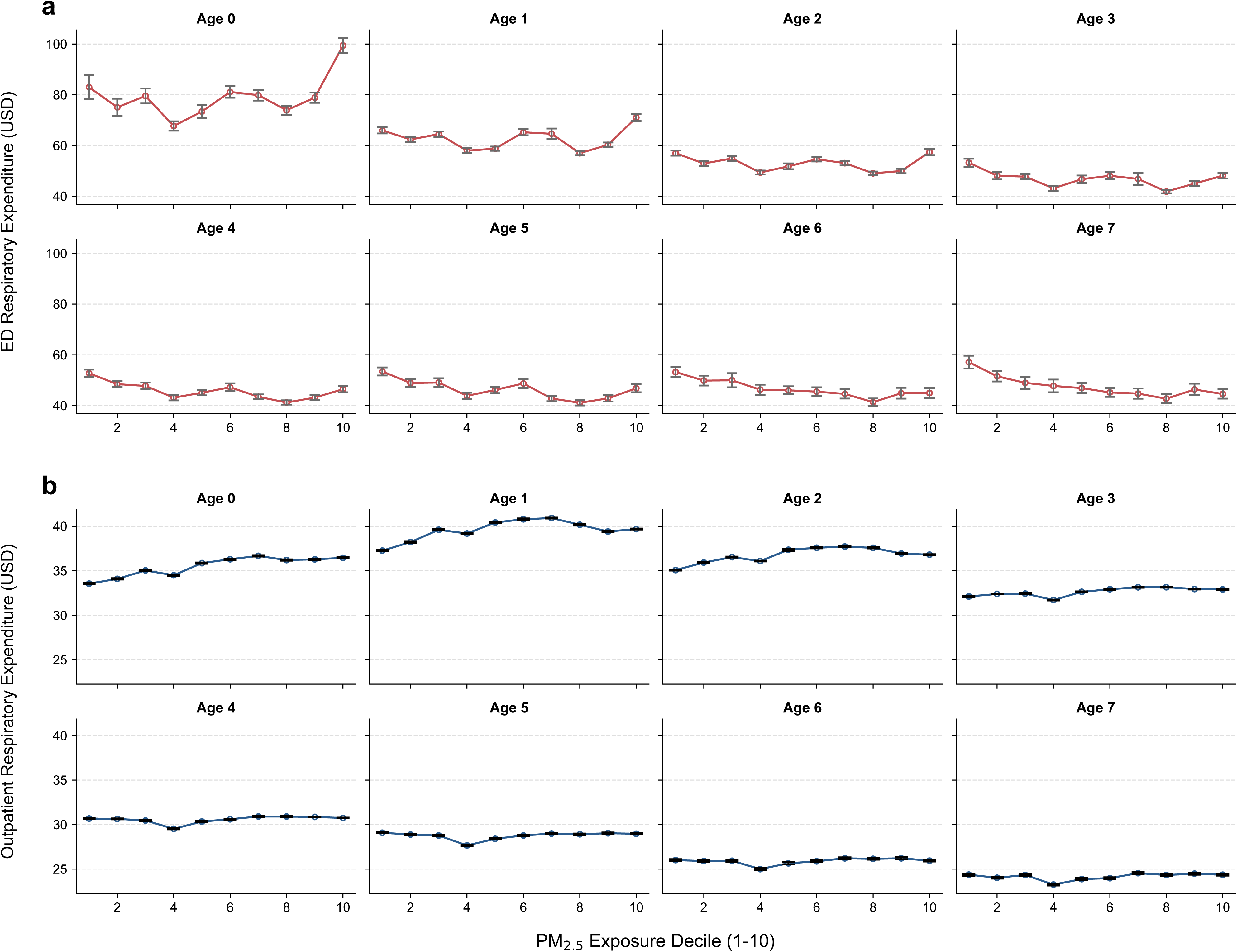
Emergency and outpatient respiratory expenditures by PM_2.5_. **a** Mean emergency department respiratory expenditure by PM_2.5_ decile (1–10) and age cohort (0–7 years). In sharp contrast to the stable outpatient trends, ED expenditures demonstrate a dramatic, non-linear spike at ages 0 and 1, specifically at the highest PM_2.5_ exposure levels (deciles 8–10). **b** Mean outpatient respiratory expenditure by PM_2.5_ decile and age cohort. Outpatient spending generally exhibits a positive association with higher pollution levels, particularly across the youngest cohorts (ages 0–2). Notably, the absolute level of outpatient spending is lower for infants aged 0 compared to those aged 1, reflecting the behavioural shielding effect during early infancy when infants are kept primarily indoors. Together, these panels provide empirical evidence that, while pollution broadly increases routine outpatient visits, the severe, high-cost clinical episodes requiring intensive resources are uniquely concentrated in the youngest infants during acute pollution events. ED emergency department.

These results reveal a dual burden: pollution broadly increases healthcare utilisation across childhood, but its financial and clinical consequences are concentrated among the youngest children. A 1-standard-deviation increase in monthly PM_2.5_ raised annual respiratory spending among children under age 5 by 32.2%. Under a simple extrapolation applying our age-specific estimates to observed spending, this corresponds to US$215 million in excess costs, driven overwhelmingly by children in their first 3 years of life (Fig. 3).

### Robustness

Placebo regression models incorporating injury and burn diagnoses produced null estimates across most age groups (Supplementary Table 3), supporting the validity of the identification strategy. Replacing mean monthly PM_2.5_ with the monthly cumulative hours exceeding 50 µg/m³ yielded closely aligned coefficients (Supplementary Table 4). Qualitatively similar age-specific patterns were observed in the post-pandemic period (2020–2023), although first-stage F statistics substantially weakened due to reduced PM_2.5_ variability, precluding strict causal inference for that subperiod (Supplementary Table 5).

## Discussion

Our findings reveal a dual burden of air-pollution exposure during childhood. While transient increases in ambient PM_2.5_ broadly elevate respiratory healthcare encounters from birth through early school age, the associated clinical and economic burdens, reflected in higher medical expenditures, are heavily concentrated among children under age 3. This divergence underscores that exposure during early development has disproportionate health and resource implications. By decomposing total respiratory expenditures, we show that this concentration is driven not by uniform increases in routine care but by a shift towards resource-intensive clinical episodes in the youngest children.

To clarify the clinical drivers of these costs, we compared expenditures across treatment intensities, contrasting routine outpatient care with high-intensity emergency encounters across PM_2.5_ deciles. Spending on high-intensity care (ED visits) is concentrated among children aged 0–1 and rises sharply at high pollution levels, whereas outpatient spending increases more uniformly across ages. This expenditure spike in the first 3 years of life when pollution is associated with a shift from milder to more clinically severe respiratory episodes, is a central empirical finding of this study. This finding aligns with the critical developmental window for rapid lung and immune maturation: physiological vulnerability in early life translates directly into economic vulnerability as pollution exposure necessitates more intensive medical interventions.

The surge in both visit frequency and ED expenditure at the transition from early infancy (0–5 months) to older infancy (6–11 months) marks a critical inflection point at which biological susceptibility meets developmental behavioural change. Immunologically, this transition coincides with the systemic depletion of passively acquired maternal immunoglobulin G, leaving older infants exceptionally vulnerable to respiratory pathogens, while PM_2.5_-induced airway inflammation acting through innate immune pathways further exacerbates this susceptibility^25–28^. Behaviourally, the 6-month milestone coincides with increasing mobility and outdoor exposure, ending the indoor shielding that characterises the first 6 months of life. Together, these biological and behavioural shifts create a developmental window of exceptional vulnerability in which ambient pollution shocks that produce only mild upper-respiratory symptoms in older children trigger high-acuity lower-respiratory emergency episodes in the youngest infants.

Scaling our age-specific estimates to observed spending reveals that routine monthly PM_2.5_ variation drives a substantial share of respiratory expenditure for children under age 5. In a universal healthcare system, higher spending per child is likely to reflect repeated episodes of severe respiratory distress. From a life-course perspective, these early-life insults may have lasting consequences: severe respiratory illness in early childhood is associated with reduced peak lung function and an increased risk of chronic obstructive pulmonary disease in adulthood, potentially limiting future health capital^38–43^. Beyond the healthcare sector, such early-life health shocks are associated with reduced educational attainment and labour-market productivity^15,44,45^. These consequences may fall disproportionately on already disadvantaged children, raising equity concerns even within a universal-coverage system^46^. As these long-term pathways are not captured by contemporaneous claims, our expenditure estimates likely represent only a fraction of pollution’s true social cost.

The weight we place on these implications depends on the credibility of the underlying causal design, which several features of our analysis support. The instrument was strong in every age group (first-stage Kleibergen–Paap F > 37), and placebo analyses using injury and burn diagnoses, which are captured through the same claims system but have no plausible physiological link to short-term particulate exposure, produced null estimates, arguing against residual confounding from general healthcare-seeking behaviour. The age gradient was also robust to replacing mean monthly PM_2.5_ with the cumulative monthly hours above 50 µg/m³, indicating that our conclusions do not depend on how exposure is operationalised. The morbidity response was of comparable magnitude in boys and girls, consistent with a shared biological susceptibility rather than sex-specific behaviour. Reassuringly, separate analyses of the post-pandemic period (2020–2023) reproduced the same age-specific pattern, indicating that the concentration of clinical severity among the youngest children is not an artefact of the study window.

This study has limitations. The principal constraint is that, although child fixed effects absorb time-invariant individual characteristics, including relatively stable differences such as whether a child is usually cared for at home or in a group setting, our administrative data cannot observe how much time each child actually spends outdoors, and therefore cannot capture individual-level personal exposure to ambient PM_2.5_. Since time spent outdoors is the proximate channel through which ambient concentrations translate into inhaled doses, this unobserved within-age variation in outdoor activity is the main source of exposure misclassification in our design. Therefore, our estimates should be interpreted as effects of ambient PM_2.5_ rather than of directly measured personal exposure. Furthermore, our estimates identify the health consequences of short-term variation in PM_2.5_ and may not extrapolate directly to sustained long-term exposure reductions. The study setting, a high-income country with universal coverage and high baseline PM_2.5_, may limit generalisability to settings with different pollution profiles or healthcare access.

These findings suggest that the burden of air pollution may be underestimated when assessed using visit counts alone, and that existing studies and regulatory assessments relying on average paediatric effects may overlook the disproportionate burden borne by the youngest children. While broad policies such as emissions standards remain essential, they should be complemented by targeted protection. As this burden is concentrated in children under age 3, targeted measures such as high-efficiency particulate air filtration in daycare centres and nurseries, or real-time air-quality alerts integrated into childcare protocols, may help reduce exposure in high-risk settings. However, their effectiveness would need to be evaluated directly. As the biological vulnerabilities underlying these patterns are not specific to Korea, the same age-targeted logic is likely to apply to other populations exposed to high ambient particulate levels. Protecting children under age 3 may therefore be particularly important for safeguarding early-life health and long-term human capital development.

## Methods

### Ethics

The study was approved by the institutional review board of Severance Hospital, Yonsei University Health System (approval number 4-2026-0583). Informed consent was waived because the analysis used de-identified administrative data. The study was conducted in accordance with the Declaration of Helsinki.

### Data construction and study population

We linked two population-scale administrative datasets from South Korea to construct a longitudinal cohort study of early childhood. The primary data source was the National Health Insurance Service database, which covers approximately 97% of the Korean population using a single-payer universal system^47^. We extracted all electronic claim records for medical services between 1 January 2015 and 31 December 2019, aggregating these daily records to the monthly level for each child (extended to 2023 for robustness checks). Each claim includes the date of service, primary and secondary diagnostic codes (ICD-10), type of facility (clinic, hospital, general hospital, or tertiary hospital), and detailed billing amounts separated into insurer payments and patient coinsurance.

The study period was strictly defined to ensure data reliability and exclude major confounding factors. The start date aligned with the nationwide standardisation of PM_2.5_ monitoring, ensuring the availability of consistent high-resolution air quality data. We excluded data from 2020 onwards from our main analysis to circumvent the confounding effects of the COVID-19 pandemic. The implementation of non-pharmaceutical interventions, most notably universal mask wearing, considerably altered paediatric healthcare utilisation patterns and the transmission dynamics of respiratory viruses^48,49^. The absolute concentration of ambient PM_2.5_ decreased considerably from 2020 onwards.

This overall reduction in pollution levels weakened the predictive power of our instrument, resulting in low first-stage F-statistics that precluded strict instrumental variable estimation for the 2020–2023 period. However, separate analyses of this later period yielded qualitatively similar age-specific patterns for both visits and expenditures (Supplementary Table 5). This suggests that while the pandemic and lower pollution levels complicated precise causal inference, the core relationship between PM_2.5_ and early childhood health remained robust even under these altered conditions.

To precisely determine the age and adjust for baseline health endowments, we merged the claims data with the National Health Screening Programme for Infants and Children (NHSPIC) using encrypted resident registration numbers. The NHSPIC is a comprehensive government-led initiative that provides eight rounds of standardised health examinations for all children from 14 days to 71 months of age. With a nationwide participation rate exceeding 90%, this programme offers near-census coverage of the birth cohort, including evaluations of physical growth, nutritional status, and developmental milestones. We restricted our sample to children born in Korea who underwent at least one screening to ensure the availability of birth weight and exact birth date information. Children with implausible birth dates, missing sex, or inconsistent insurance eligibility were excluded. The final analytical sample consisted of a child-month panel, tracking individual children from birth until they reached 95 months of age or at the end of the study period, yielding 99,261,945 child-month observations from 3,210,381 unique children.

### Air pollution and meteorological data

To construct the thermal inversion instrument and establish a spatial framework for our analysis, we used the NASA Modern-Era Retrospective Analysis for Research and Applications dataset (M2I6NPANA, Version 5.12.4). This dataset provides global 6-hourly temperature measurements across 42 pressure levels on a 0.625° × 0.5° spatial grid. While the South Korean peninsula is covered by 45 such grid cells, we excluded 10 cells located in the high-elevation Taebaek mountains region owing to invalid surface temperature readings (missing 1000 hPa data). Consequently, the final analysis relied on the remaining 35 valid grid cells.

We calculated the thermal inversion strength by computing the vertical temperature gradient between the 1,000 and 975 hPa pressure levels at 6-hour intervals. We defined the inversion intensity based on the positive temperature difference (T975hPa − T1000hPa > 0), treating the non-inversion periods as zero. These values were averaged to generate a monthly inversion strength index for each grid cell.

For local air quality and weather controls, we obtained hourly PM_2.5_ and precipitation data from the National Institute of Environmental Research and Korea Meteorological Administration.^50^ As these datasets provide measurements from monitoring stations with precise GPS coordinates, we spatially mapped each station to one of the 35 valid grid cells defined above. Given South Korea’s exceptionally dense monitoring network (exceeding 500 stations nationwide as of 2023), grid-cell averaging provides a robust measure of local exposure and minimises potential measurement error. We then calculated the arithmetic mean of the readings from all stations within each cell to construct the monthly environmental variables. Finally, these cell-level measures were matched to administrative districts (Si-Gun-Gu) to assign local exposure.

### Outcome definitions

We defined two primary dependent variables to capture the distinct margins of healthcare utilisation. First, the morbidity margin (frequency) was defined as the total number of respiratory-related encounters per child per month. To capture the extensive margin of pollution-triggered medical attention, we restricted respiratory cases to acute infections and asthma (ICD-10: J00–J98) while excluding contagious viral epidemics (Supplementary Table 6). Second, the expenditure margin (severity/cost) represents the total monthly respiratory healthcare spending per child, combining the insurer’s reimbursement and the patient’s out-of-pocket payment. Given the frequent healthcare utilisation observed in our study population, with a monthly average of 1.71 respiratory visits per child, we focused specifically on addressing the skewness of the cost data. We defined the expenditure outcome as the natural logarithm of monthly medical expenditure plus one [log(expenditure + 1)]. This transformation accounts for the right-skewed distribution of costs and crucially allows for the inclusion of months with zero spending, capturing the unconditional financial burden across the entire population. Visit counts were analysed without transformation. All monetary values were deflated to 2020 South Korean Won (KRW) using the Consumer Price Index and converted to US dollars using a fixed exchange rate of 1,300 KRW/USD.

### Empirical strategy

To estimate the causal effect of PM_2.5_ on child health, we employed a two-stage least squares regression, exploiting thermal inversions as an exogenous shifter of local air quality. Previous studies in Korea have often used westerly winds as instruments to capture the transboundary transport of particulates from continental East Asia^32,36,37^. However, that strategy identifies only the imported component of pollution and misses domestically generated emissions and high-PM_2.5_ episodes arising under non-westerly conditions. Thermal inversions instead act on the total particulate load already present in the boundary layer, trapping domestic and transboundary emissions alike regardless of wind direction. Identification rests on the exclusion restriction that inversion-driven variation in PM_2.5_ affects paediatric respiratory outcomes only through pollution exposure, and not through other channels such as ambient temperature or co-occurring viral epidemics. Two features support this assumption. First, year and month fixed effects absorb seasonal drivers, including average seasonal temperature and influenza and respiratory syncytial virus cycles, that could otherwise accompany inversion episodes. Second, placebo analyses of injury and burn diagnoses, which are outcomes with no physiological link to short-term particulate exposure, yield null estimates (Supplementary Table 3).

The first-stage equation estimated the relationship between thermal inversions and local PM_2.5_ concentrations (equation (1)), while the second stage estimated the effect of pollution on the respiratory health burden (equation (2)):

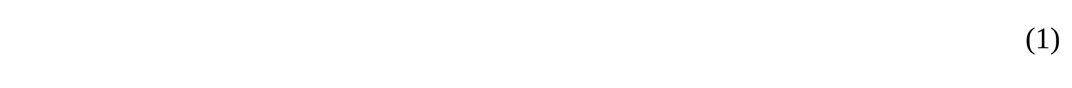

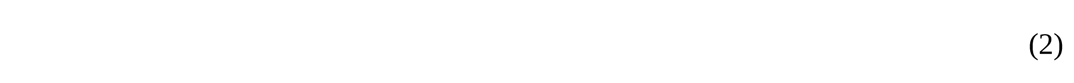

(Here, denotes the monthly health outcome for child *i* residing in grid cell *g* in month *t* (either the number of respiratory visits or log expenditure), and is the predicted monthly PM_2.5_ concentration from the first stage. Crucially, our model includes a rich set of fixed effects and controls to isolate causal signals. Child fixed effects () absorb all time-invariant child characteristics, including genetic health endowment, sex, and parental background. To control for temporal confounders, we included year and month fixed effects (), which account for long-term trends (e.g. medical inflation) and nationwide seasonality (e.g. flu cycles), respectively. The vector of time-varying controls () includes monthly precipitation to account for wash-out effects, household income (proxied by insurance premiums), parental occupation, and disability status. Standard errors were clustered at the grid-cell level to account for the serial correlation and spatial dependence of pollution errors within the environmental grid structure. All models were estimated separately for each annual age group (ages 0–7) and additionally by sex to examine heterogeneity (Supplementary Tables 1 and 2). First-stage Kleibergen–Paap F statistics exceeded 37 in all age groups.

### Robustness and placebo tests

We conducted sensitivity analyses to validate our findings. First, we stratified the samples according to sex to test for biological heterogeneity. Second, we replaced mean monthly PM_2.5_ with high-pollution hours—the monthly cumulative hours with PM_2.5_ > 50 µg/m³—to test sensitivity to acute exposure peaks. The resulting estimates are closely aligned with our main results (Supplementary Table 4). Finally, we used injuries or burns (ICD-10: S10–S99, T00–T14) as placebo outcomes. These outcomes share healthcare-seeking behaviour and are reported through the same claims system but are not physiologically linked to short-term ambient PM_2.5_ exposure. We observed no systematic association between these non-respiratory outcomes, providing further support for our identification strategy (Supplementary Table 3). The isolated signal at age 7 lacks consistency across specifications and likely reflects statistical noise rather than a biological mechanism.

### Statistics

Causal effects were estimated by two-stage least squares (2SLS) instrumental-variable regression, with thermal-inversion strength instrumenting for monthly grid-cell PM_2.5_. Standard errors were clustered at the grid-cell level, and instrument strength was confirmed by first-stage Kleibergen–Paap F statistics exceeding 37 in all age groups. Models were estimated separately for each annual age group (ages 0–7) and by sex, using the full analytical sample of 3,210,381 children and 99,261,945 child-month observations. All statistical tests were two-sided; estimates were considered statistically significant at the 5% level, inferred from whether the 95% confidence interval excluded zero. 95% confidence intervals for all coefficients, including non-significant ones, are reported in the corresponding main-text and Supplementary Tables. In all figures, error bars and shaded bands denote 95% confidence intervals. Analyses were performed in Stata 17.0 (StataCorp) and SAS 9.4 (SAS Institute).

### Reporting summary

Further information on research design is available in the Nature Portfolio Reporting Summary linked to this article.

## Data availability

Aggregate regional PM_2.5_ concentrations are publicly available from AirKorea, managed by the Korea Environment Corporation (https://www.airkorea.or.kr). Thermal-inversion data were accessed from the NASA GES DISC archive (M2I6NPANA; https://disc.gsfc.nasa.gov). Meteorological data were obtained from the Korea Meteorological Administration Open Data Portal (https://data.kma.go.kr). Individual-level NHIS and NHSPIC microdata are not publicly available owing to legal restrictions protecting patient privacy under the Personal Information Protection Act of Korea, but may be accessed for research following formal application and approval through the National Health Insurance Data Sharing Service (https://nhiss.nhis.or.kr).

## Code availability

The complete analytical code for data extraction, variable construction, and statistical modelling is available at Zenodo (https://doi.org/10.5281/zenodo.19953320) under a Creative Commons Attribution licence. SAS and Stata were used for the analyses.

## Acknowledgements

This work was supported by the Ministry of Education of the Republic of Korea and the National Research Foundation of Korea (NRF-2025S1A5B5A17018793).

## Author contributions

H.K. and H.-S.Y. conceptualised the study and designed the methodology. H.K. and J.Shin conducted data processing and statistical analysis. J.Suh provided clinical interpretation of paediatric respiratory outcomes and emergency care. H.K. drafted the manuscript. H.-S.Y. supervised the project. H.K. and J.Shin accessed and verified the underlying data. All authors had full access to all the data and approved the submitted manuscript.

## Additional information

Supplementary Information is available for this paper.

Correspondence and requests for materials should be addressed to Jaeyong Shin or Hee-Seung Yang.

## Supplementary Tables

**Supplementary Table 1.** Age-specific causal effects of PM_2.5_ on respiratory outcomes (boys, 2015–2019)

| Outcome variable | Age 0 | Age 1 | Age 2 | Age 3 | Age 4 | Age 5 | Age 6 | Age 7 |
| --- | --- | --- | --- | --- | --- | --- | --- | --- |
| **Panel A. Respiratory visits (count per month)** |  |  |  |  |  |  |  |  |
| PM<sub>2.5</sub> | 0.027<br>(0.009–0.046) | 0.041<br>(0.011–0.071) | 0.034<br>(0.014–0.055) | 0.020<br>(0.007–0.033) | 0.013<br>(0.004–0.023) | 0.010<br>(0.001–0.019) | 0.007<br>(0.000–0.015) | 0.004<br>(–0.003–0.011) |
| P-value | 0.003 | 0.007 | 0.001 | 0.003 | 0.006 | 0.028 | 0.046 | 0.286 |
| **Panel B. Respiratory expenditure (log, US\$)** | | | | | | | | |
| PM<sub>2.5</sub> | 0.053<br>(0.011–0.095) | 0.048<br>(0.003–0.092) | 0.043<br>(0.013–0.073) | 0.025<br>(0.002–0.049) | 0.019<br>(–0.001–0.040) | 0.021<br>(–0.004–0.047) | 0.021<br>(–0.003–0.044) | 0.009<br>(–0.023–0.041) |
| P-value | 0.014 | 0.035 | 0.005 | 0.034 | 0.064 | 0.102 | 0.086 | 0.577 |
| Income | Y | Y | Y | Y | Y | Y | Y | Y |
| Precipitation | Y | Y | Y | Y | Y | Y | Y | Y |
| Parental occupation | Y | Y | Y | Y | Y | Y | Y | Y |
| Disability status | Y | Y | Y | Y | Y | Y | Y | Y |
| Individual FE | Y | Y | Y | Y | Y | Y | Y | Y |
| Year & month FE | Y | Y | Y | Y | Y | Y | Y | Y |
| KP F statistic | 36.77 | 39.52 | 38.85 | 42.37 | 41.43 | 47.96 | 55.27 | 51.76 |
| Unique IDs | 631,802 | 653,801 | 696,290 | 714,855 | 714,354 | 697,599 | 687,500 | 681,933 |
| Observations | 5,726,241 | 5,869,661 | 6,296,543 | 6,474,837 | 6,472,083 | 6,359,275 | 6,265,323 | 6,147,922 |

Notes: Second-stage regression results for boys using data from 2015 to 2019, excluding the COVID-19 period. Panel A reports estimates for monthly respiratory visits per child-month, and Panel B reports estimates for log monthly respiratory healthcare expenditure per child-month. Columns ‘Age 0’ through ‘Age 7’ correspond to children aged 0–11, 12–23, 24–35, 36–47, 48–59, 60–71, 72–83, and 84–95 months, respectively. All models include child, year, and month fixed effects and controls for precipitation, household income, parental occupation, and disability status. Effect sizes are 2SLS coefficients; 95% confidence intervals, computed from standard errors clustered at the grid-cell level, are reported in parentheses beneath each coefficient. 2SLS two-stage least squares, CI confidence interval, FE fixed effects, KP Kleibergen–Paap.

**Supplementary Table 2.** Age-specific causal effects of PM_2.5_ on respiratory outcomes (girls, 2015–2019)

| Outcome variable | Age 0 | Age 1 | Age 2 | Age 3 | Age 4 | Age 5 | Age 6 | Age 7 |
| --- | --- | --- | --- | --- | --- | --- | --- | --- |
| **Panel A. Respiratory visits (count per month)** |  |  |  |  |  |  |  |  |
| PM<sub>2.5</sub> | 0.025<br>(0.007–0.043) | 0.040<br>(0.011–0.069) | 0.035<br>(0.014–0.056) | 0.022<br>(0.009–0.035) | 0.013<br>(0.004–0.021) | 0.010<br>(0.002–0.018) | 0.008<br>(0.000–0.016) | 0.005<br>(–0.002–0.011) |
| P-value | 0.006 | 0.007 | 0.001 | 0.001 | 0.003 | 0.018 | 0.051 | 0.173 |
| **Panel B. Respiratory expenditure (log, US\$)** | | | | | | | | |
| PM<sub>2.5</sub> | 0.053<br>(0.009–0.096) | 0.051<br>(0.007–0.096) | 0.041<br>(0.010–0.072) | 0.031<br>(0.007–0.055) | 0.016<br>(–0.003–0.035) | 0.020<br>(–0.002–0.041) | 0.027<br>(–0.001–0.056) | 0.017<br>(–0.010–0.044) |
| P-value | 0.017 | 0.024 | 0.009 | 0.011 | 0.090 | 0.077 | 0.062 | 0.212 |
| Income | Y | Y | Y | Y | Y | Y | Y | Y |
| Precipitation | Y | Y | Y | Y | Y | Y | Y | Y |
| Parental occupation | Y | Y | Y | Y | Y | Y | Y | Y |
| Disability status | Y | Y | Y | Y | Y | Y | Y | Y |
| Individual FE | Y | Y | Y | Y | Y | Y | Y | Y |
| Year & month FE | Y | Y | Y | Y | Y | Y | Y | Y |
| KP F statistic | 37.29 | 40.26 | 40.06 | 42.16 | 43.70 | 47.05 | 54.95 | 51.07 |
| Unique IDs | 598,089 | 619,414 | 659,500 | 677,730 | 675,567 | 658,088 | 648,197 | 642,350 |
| Observations | 5,421,494 | 5,559,546 | 5,971,667 | 6,141,380 | 6,120,538 | 5,995,918 | 5,909,195 | 5,785,718 |

Notes: Second-stage regression results for girls using data from 2015 to 2019, excluding the COVID-19 period. Panel A reports estimates for monthly respiratory visits per child-month, and Panel B reports estimates for log monthly respiratory healthcare expenditure per child-month. Columns ‘Age 0’ through ‘Age 7’ correspond to children aged 0–11, 12–23, 24–35, 36–47, 48–59, 60–71, 72–83, and 84–95 months, respectively. All models include child, year, and month fixed effects and controls for precipitation, household income, parental occupation, and disability status. Effect sizes are 2SLS coefficients; 95% confidence intervals, computed from standard errors clustered at the grid-cell level, are reported in parentheses beneath each coefficient. 2SLS two-stage least squares, CI confidence interval, FE fixed effects, KP Kleibergen–Paap.

**Supplementary Table 3.** Causal effects of PM_2.5_ on injuries and burns (2015–2019)

| Outcome variable | Age 0 | Age 1 | Age 2 | Age 3 | Age 4 | Age 5 | Age 6 | Age 7 |
| --- | --- | --- | --- | --- | --- | --- | --- | --- |
| **Panel A. Injury and burn visits (count per month)** |  |  |  |  |  |  |  |  |
| PM<sub>2.5</sub> | 0.0002<br>(-0.0002–<br>0.0005) | 0.0004<br>(0.0000–<br>0.0007) | 0.0001<br>(-0.0007–<br>0.0009) | 0.0003<br>(-0.0004–<br>0.0010) | 0.0003<br>(-0.0001–<br>0.0006) | 0.0001<br>(-0.0009–<br>0.0010) | 0.0001<br>(-0.0005–<br>0.0006) | 0.0010<br>(0.0006–<br>0.0014) |
| P-value | 0.420 | 0.072 | 0.835 | 0.397 | 0.132 | 0.882 | 0.760 | <0.001 |
| **Panel B. Injury and burn expenditure (log, US\$)** | | | | | | | | |
| PM<sub>2.5</sub> | 0.0003<br>(-0.0014–<br>0.0021) | 0.0017<br>(-0.0002–<br>0.0037) | 0.0001<br>(-0.0060–<br>0.0063) | 0.0012<br>(-0.0017–<br>0.0041) | 0.0013<br>(-0.0006–<br>0.0032) | -0.0001<br>(-0.0036–<br>0.0033) | -0.0001<br>(-0.0017–<br>0.0015) | 0.0024<br>(0.0005–<br>0.0042) |
| P-value | 0.705 | 0.081 | 0.962 | 0.428 | 0.191 | 0.945 | 0.884 | 0.012 |
| Income | Y | Y | Y | Y | Y | Y | Y | Y |
| Precipitation | Y | Y | Y | Y | Y | Y | Y | Y |
| Parental occupation | Y | Y | Y | Y | Y | Y | Y | Y |
| Disability status | Y | Y | Y | Y | Y | Y | Y | Y |
| Individual FE | Y | Y | Y | Y | Y | Y | Y | Y |
| Year & month FE | Y | Y | Y | Y | Y | Y | Y | Y |
| KP F statistic | 31.37 | 36.18 | 36.74 | 40.44 | 41.22 | 46.99 | 55.68 | 51.86 |
| Unique IDs | 827,918 | 918,273 | 1,038,035 | 1,120,780 | 1,167,116 | 1,177,763 | 1,190,688 | 1,202,546 |
| Observations | 7,522,315 | 8,265,762 | 9,413,175 | 10,181,493 | 10,609,032 | 10,766,700 | 10,880,617 | 10,864,374 |

Notes: Second-stage regression results using data from 2015 to 2019, excluding the COVID-19 period. Panel A reports estimates for monthly healthcare visits for injuries or burns (ICD-10 S10–S99, T00–T14) per child-month, and Panel B reports estimates for log monthly healthcare expenditure for those conditions per child-month. Columns ‘Age 0’ through ‘Age 7’ correspond to children aged 0–11, 12–23, 24–35, 36–47, 48–59, 60–71, 72–83, and 84–95 months, respectively. All models include child, year, and month fixed effects and controls for precipitation, household income, parental occupation, and disability status. Effect sizes are 2SLS coefficients; 95% confidence intervals, computed from standard errors clustered at the grid-cell level, are reported in parentheses beneath each coefficient. 2SLS two-stage least squares, CI confidence interval, FE fixed effects, KP Kleibergen–Paap. ICD-10 International Classification of Diseases 10th revision.

**Supplementary Table 4.** Alternative metric: high-pollution hours (PM_2.5_ > 50 µg/m³)

| Outcome variable | Age 0 | Age 1 | Age 2 | Age 3 | Age 4 | Age 5 | Age 6 | Age 7 |
| --- | --- | --- | --- | --- | --- | --- | --- | --- |
| **Panel A. Respiratory visits (count per month)** |  |  |  |  |  |  |  |  |
| High PM<sub>2.5</sub> | 0.0019<br>(0.0006–<br>0.0032) | 0.0029<br>(0.0008–<br>0.0049) | 0.0024<br>(0.0010–<br>0.0039) | 0.0015<br>(0.0006–<br>0.0024) | 0.0009<br>(0.0003–<br>0.0015) | 0.0007<br>(0.0001–<br>0.0013) | 0.0005<br>(0.0000–<br>0.0010) | 0.0003<br>(–0.0002–<br>0.0008) |
| P-value | 0.005 | 0.006 | 0.001 | 0.002 | 0.003 | 0.021 | 0.038 | 0.234 |
| **Panel B. Respiratory expenditure (log, US\$)** | | | | | | | | |
| High PM<sub>2.5</sub> | 0.0038<br>(0.0007–<br>0.0070) | 0.0036<br>(0.0005–<br>0.0067) | 0.0030<br>(0.0009–<br>0.0052) | 0.0021<br>(0.0004–<br>0.0037) | 0.0013<br>(0.0000–<br>0.0026) | 0.0014<br>(–0.0002–<br>0.0030) | 0.0017<br>(–0.0001–<br>0.0035) | 0.0009<br>(–0.0011–<br>0.0030) |
| P-value | 0.018 | 0.025 | 0.006 | 0.015 | 0.055 | 0.084 | 0.062 | 0.381 |
| Income | Y | Y | Y | Y | Y | Y | Y | Y |
| Precipitation | Y | Y | Y | Y | Y | Y | Y | Y |
| Parental occupation | Y | Y | Y | Y | Y | Y | Y | Y |
| Disability status | Y | Y | Y | Y | Y | Y | Y | Y |
| Individual FE | Y | Y | Y | Y | Y | Y | Y | Y |
| Year & month FE | Y | Y | Y | Y | Y | Y | Y | Y |
| KP F statistic | 43.75 | 53.32 | 50.30 | 54.79 | 57.39 | 60.32 | 69.12 | 59.39 |
| Unique IDs | 1,241,727 | 1,284,526 | 1,367,750 | 1,404,185 | 1,400,523 | 1,365,847 | 1,345,869 | 1,334,600 |
| Observations | 11,268,090 | 11,545,088 | 12,390,134 | 12,733,622 | 12,703,393 | 12,458,849 | 12,276,575 | 12,041,087 |

Notes: Second-stage estimates using the monthly count of hours in which PM<sub>2.5</sub> concentrations exceeded 50 µg/m<sup>3</sup> as the exposure variable, based on data from 2015 to 2019, excluding the COVID-19 period. Panel A reports estimates for monthly respiratory visits per child-month, and Panel B reports estimates for log monthly respiratory healthcare expenditure per child-month. Columns ‘Age 0’ through ‘Age 7’ correspond to children aged 0–11, 12–23, 24–35, 36–47, 48–59, 60–71, 72–83, and 84–95 months, respectively. All models include child, year, and month fixed effects and controls for precipitation, household income, parental occupation, and disability status. Effect sizes are 2SLS coefficients; 95% confidence intervals, computed from standard errors clustered at the grid-cell level, are reported in parentheses beneath each coefficient. 2SLS two-stage least squares, CI confidence interval, FE fixed effects, KP Kleibergen–Paap.

**Supplementary Table 5.** Age-specific causal effects of PM_2.5_ on respiratory outcomes (2020–2023)

| Outcome variable | Age 0 | Age 1 | Age 2 | Age 3 | Age 4 | Age 5 | Age 6 | Age 7 |
| --- | --- | --- | --- | --- | --- | --- | --- | --- |
| **Panel A. Respiratory visits (count per month)** |  |  |  |  |  |  |  |  |
| PM<sub>2.5</sub> | 0.030<br>(0.004–0.056) | 0.099<br>(0.022–0.176) | 0.131<br>(0.032–0.229) | 0.114<br>(0.033–0.196) | 0.070<br>(0.007–0.132) | 0.030<br>(–0.039–0.099) | –0.008<br>(–0.074–0.058) | –0.012<br>(–0.073–0.049) |
| P-value | 0.024 | 0.012 | 0.009 | 0.006 | 0.030 | 0.394 | 0.810 | 0.704 |
| **Panel B. Respiratory expenditure (log, US\$)** | | | | | | | | |
| PM<sub>2.5</sub> | 0.070<br>(–0.035–0.176) | 0.201<br>(0.010–0.393) | 0.284<br>(0.066–0.502) | 0.272<br>(0.077–0.466) | 0.185<br>(–0.028–0.399) | 0.099<br>(–0.157–0.354) | –0.014<br>(–0.284–0.256) | –0.029<br>(–0.305–0.248) |
| P-value | 0.191 | 0.039 | 0.011 | 0.006 | 0.089 | 0.449 | 0.920 | 0.839 |
| Income | Y | Y | Y | Y | Y | Y | Y | Y |
| Precipitation | Y | Y | Y | Y | Y | Y | Y | Y |
| Parental occupation | Y | Y | Y | Y | Y | Y | Y | Y |
| Disability status | Y | Y | Y | Y | Y | Y | Y | Y |
| Individual FE | Y | Y | Y | Y | Y | Y | Y | Y |
| Year & month FE | Y | Y | Y | Y | Y | Y | Y | Y |
| KP F statistic | 2.71 | 2.79 | 2.90 | 2.83 | 2.85 | 2.88 | 2.75 | 2.69 |
| Unique IDs | 772,640 | 856,457 | 932,428 | 1,000,345 | 1,075,725 | 1,126,623 | 1,158,320 | 1,190,552 |
| Observations | 6,924,702 | 7,256,575 | 8,047,767 | 8,664,581 | 9,361,478 | 9,929,422 | 10,255,525 | 10,413,470 |

Notes: Second-stage regression results using data from 2020 to 2023, covering the post-pandemic period. Panel A reports estimates for monthly respiratory visits per child-month, and Panel B reports estimates for log monthly respiratory healthcare expenditure per child-month. Columns ‘Age 0’ through ‘Age 7’ correspond to children aged 0–11, 12–23, 24–35, 36–47, 48–59, 60–71, 72–83, and 84–95 months, respectively. Low first-stage F statistics in this period reflect pandemic-induced disruptions in healthcare utilisation and air pollution patterns and preclude strict instrumental-variable inference for this subperiod. All models include child, year, and month fixed effects and controls for precipitation, household income, parental occupation, and disability status. Effect sizes are 2SLS coefficients; 95% confidence intervals, computed from standard errors clustered at the grid-cell level, are reported in parentheses beneath each coefficient. 2SLS two-stage least squares, CI confidence interval, FE fixed effects, KP Kleibergen–Paap.

**Supplementary Table 6.** ICD-10 codes for respiratory outcomes.

| Category | ICD-10 codes | Included conditions |
| --- | --- | --- |
| Acute upper infections | J00–J04, J06 | Nasopharyngitis, pharyngitis, tonsillitis |
| Pneumonia | J12–J15, J18 | Viral and bacterial pneumonia |
| Acute lower infections | J20–J22 | Bronchitis and bronchiolitis |
| Other upper diseases | J30–J32 | Allergic rhinitis and sinusitis |
| Chronic lower diseases | J40–J42, J45–J46 | Asthma and bronchitis |
| Other disorders | J68–J69, J77, J80, J96, J98 | ARDS, respiratory failure, etc. |

Notes: To isolate the acute environmental shock of PM<sub>2.5</sub>, we a priori restricted outcomes to biologically plausible paediatric responses, explicitly excluding confounders such as contagious viral epidemics (J05, J09–J11), adult-onset morbidities (J43–J44), and anatomical abnormalities (J33). ICD-10 International Classification of Diseases 10th revision.

